# Basic reproduction number projection for novel pandemics and variants Ancestral SARS-CoV2 ℛ_0_ projection

**DOI:** 10.1101/2022.07.03.22277191

**Authors:** Ryan L. Benjamin

## Abstract

The recently derived Hybrid-Incidence Susceptible-Transmissible-Removed (HI-STR) prototype is a deterministic epidemic compartment model and an alternative to the Susceptible-Infected-Removed (SIR) model prototype. The HI-STR predicts that pathogen transmission depends on host population characteristics including population size, population density and some common host behavioural characteristics.

The HI-STR prototype is applied to the ancestral Severe Acute Respiratory Syndrome Coronavirus 2 (SARS-CoV2) to show that the original estimates of the Coronavirus Disease 2019 (COVID-19) basic reproduction number (*ℛ*_0_) for the United Kingdom (UK) could have been projected on the individual states of the United States of America (USA) prior to being detected in the USA.

The Imperial College London (ICL) group’s *ℛ*_0_ estimate for the UK is projected onto each USA state. The difference between these projections and ICL’s estimates for USA states is either not statistically significant on the paired student t-test or epidemiologically insignificant.

Projection provides a baseline for evaluating the real-time impact of an intervention. Sensitivity analysis was conducted because of considerable variance in parameter estimates across studies. Although the HI-STR predicts that increasing symptomatic ratio and inherently immune ratio reduce *ℛ*_0_, relative to the uncertainty in the estimates of *ℛ*_0_ for the ancestral SARS-CoV2, the projection is insensitive to the inherently immune ratio and the symptomatic ratio.

## 1 Introduction

### 1.1 Motivation

The coronavirus disease 2019 (COVID-19) pandemic highlighted the need to anticipate the impact of a novel pathogen on healthcare [1–4] or the economy [5, 6]. One of the impact factors is the basic reproduction number (*ℛ*_0_) – a demographic concept that has been repurposed for infectious disease epidemiology [7–11]. *ℛ*_0_ represents the average number of susceptible people a host infects in a completely susceptible population whilst that host is in its infected state [12, 13]. Based on *ℛ*_0_ estimates for COVID19’s causative agent, severe acute respiratory syndrome coronavirus 2 (SARS-CoV2), various categories of predictive [1, 14–17], forecast [18–20] and regression [21–24] models have been constructed to anticipate healthcare system demand.

The COVID-19 pandemic’s infections have been periodic [23–27]. Continuous feedback control loops like prevalence dependent contact rates [28] and intervention fatigue [29] may contribute. Equally, irregular events/pulses like relaxation of previous restrictions, superspreader events and migration [29] result in perturbations in the rate of new infections or the active infections [25]. Cyclical events like seasonal host behaviour or pathogen biology, seasonal migration or waning immunity [27, 29] result in periodic infection perturbations or vibrations [24]. The superposition of these perturbations manifest as pandemic waves [30].

For the COVID-19 pandemic, some subsequent waves of infection have been associated with mutations to the ancestral (wild-type) SARS-CoV2 in some countries [29, 31, 32]. Paradoxically, these distinct variant waves may be consequence of SARS-CoV2’s slow virion mutation rate [33–35]. Even if the virion mutation rate is constant, the time to accumulate the appropriate number of mutations in the appropriate loci of a virion, in a sufficiently gregarious individual, in a sufficiently connected geographical location to collectively constitute a variant of concern (VOC) may not be [35]. Thus the timing and the impact of these these events are treated as random and prediction requires manifestation in at least one region. This manuscript projects the impact of a random event like a novel VOC from a region in which it has manifested to one in which it has not. It proposes that each of SARS-CoV2’s VOCs (with its associated perturbations) can be treated as a pandemic and that COVID-19 is the collective manifestations of these overlapping pandemics [36]. Their distinct clinical manifestations provudes justification for this approach [37,38].

Implicitly each VOC contender is a potential new pandemic [29, 34, 39–41]. Consequently, a new local *ℛ*_0_ can be projected for that VOC. This local *ℛ*_0_ represents an upperbound of the challenger VOC’s impact in anticipation of it outcompeting and supplanting the incumbent [42]. It is an upperbound because, by definition, a *ℛ*_0_ assumes complete susceptibility to the new variant.

The hybrid-incidence, susceptible-transmissible-removed (HI-STR) [43] model is a deterministic, compartment model prototype constructed to replace two assumptions of Kermack-McKendrick’s susceptible-infectious-removed (SIR) prototype [44–46]. It replaces the assumption that the removal rate is proportional to the size of the infected compartment with the more biologically appropriate assumption that the transmissible period is fixed and, consequently, the removal rate is the same as the infection rate one transmissible period ago [43]. It also replaces Hamer’s mass action law with its chemistry precursor – the law of mass action [47]. The latter allows the derivation of a population density dependent *R*_0_ [21, 22, 43].

The HI-STR model differs from existing compartment models by predicting that *ℛ*_0_ is not only a pathogen property but also depends on the host population’s characteristics – including population-size (*N*), -density (*ρ*_*n*_) and behaviour [29, 48]. Here a novel method of foretelling local ℛ_0_ in sufficiently behaviourally-similar, isolated populations is introduced. This method is des-similar ignated projection. It proposes that, if an estimate of *ℛ*_0_ exists for an isolated population 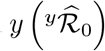, the projection of this *ℛ*_0_ onto a sufficiently behaviourally similar isolated population *z* is given by

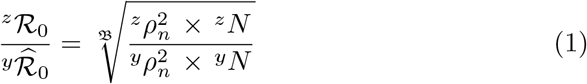

Where **ℬ** is determined by the pathogen’s transmission dynamics in that host population.

### 1.2 Background

The omnipresent SIR compartment model prototype for the temporal evolution of an infectious disease proposes that the individuals of an homogenous population can be grouped into three compartments – susceptible, infected and removed [44–46]. Susceptible implies capable of contracting a pathogen, infected implies capable of replicating and spreading the pathogen and removed refers to either recovery (expulsion of the pathogen and immunity) or death. Additional compartments [1, 49] and stratified or heterogenous populations [50–53] result in more sophisticated deterministic, compartment models.

An infectious epidemiology modelling taxonomy is proposed (Figure 1) to distinguish between foretell’s common synonyms [54] in mathematical epidemiology. It is proposed that deterministic, compartment models are a subcategory of differential equation (DE), orthodox, predictive models. Predictive (mechanistic [55]) models presuppose that phenomena can be explained and that these explanations can be simulated. The orthodox predictive models, consist of a three or four step process of explanation, abstraction into mathematics, the application of a numerical method and *in silico* simulation of the abstraction. The pioneering categories of orthodox models are stochastic and deterministic.

**Fig. 1.**
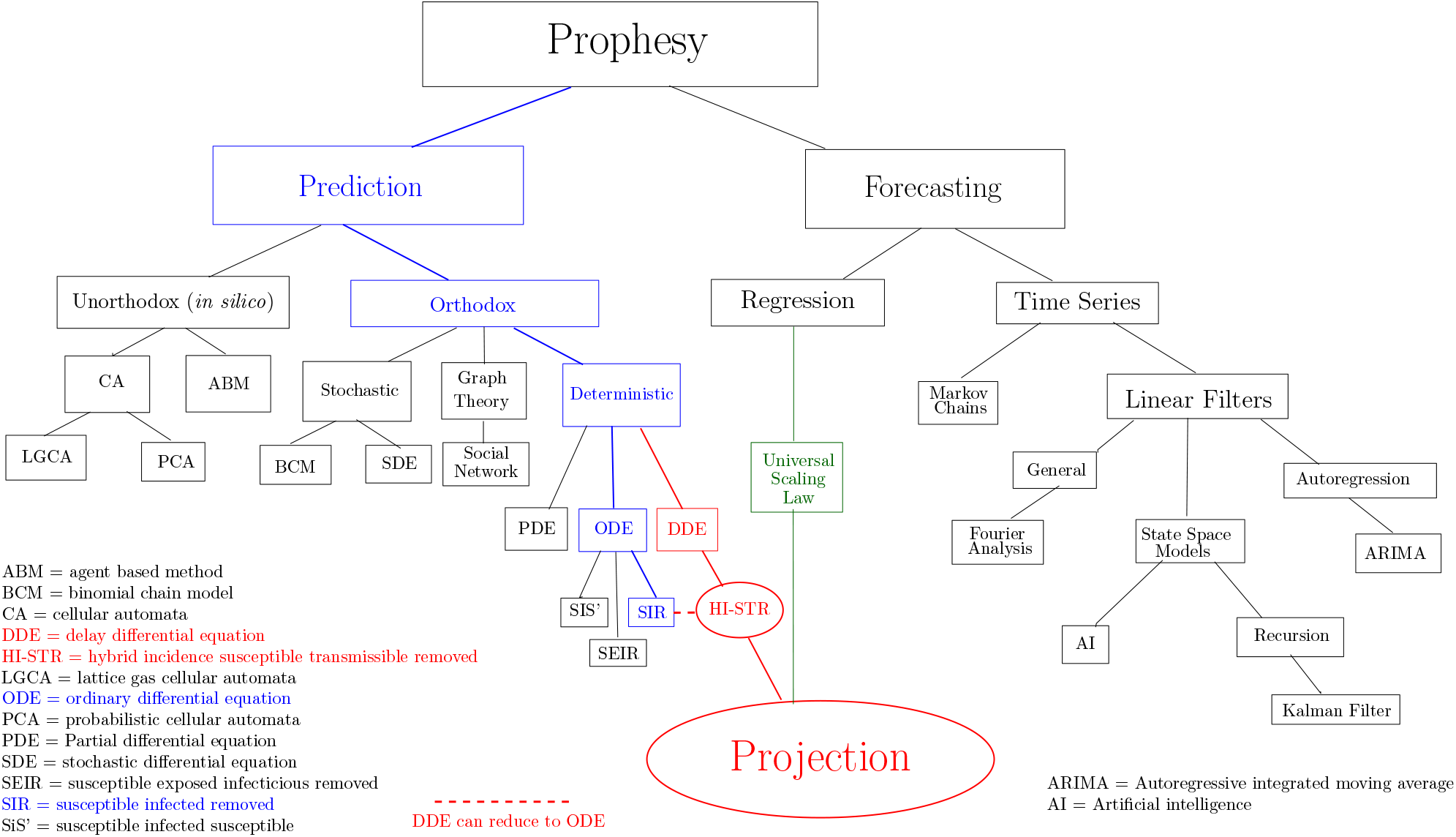
Epidemiology prophesy taxonomy

The deterministic compartment models are DE models. The DEs assume an homogenous population and simulate averaged phenomena. The ordinary differential equation (ODE) models only simulate the rate of change of the compartment sizes. Historically, the delay differential equation (DDE) compartment models [56, 57] are an alternative to the exposed (E) compartment of the SEIR ODE model [58–60]. Both the delay term and the E compartment incorporate an incubation period into the SIR prototype. The HI-STR is a DDE model that reduces to an ODE for periodic phenomena [43]. The HI-STR’s delay is not due to incubation, it is intended to simulate a constant transmissible period. The transmissible period is another subtle difference from conventional ODE models. Similar to the infectious period, it is the period of time that a host can transmit the disease but it can be limited biologically (*e*.*g*. the incubation period), behaviourally (*e*.*g*. isolation, quarantine [61] or hospitalisation) or technologically (*e*.*g*. face mask or pharmacy). An example of pharmacological restriction to a transmissible period is human immunodeficiency virus (HIV) control where anti-retrovirals (ARVs) substantially reduce viral load and therefore transmissibility. Thus transmissibility may be idealised as a step function under appropriate circumstances [62]. Implicitly, transmissibility is a population characteristic while infectivity is an individual characteristic. Partial differential equation (PDE) models typically model spatial spread as diffusion [63–65]. Algebraic formulae for thresholds like _0_ and proportion to vaccinate are a consequence of deterministic models

Stochastic, orthodox models translate to binomial chain models (BCM) [66–68] or stochastic differential equation (SDE) models that superimpose uncertainty on ODE models [69–71]. They complement deterministic models with their ability to assign probabilities to outlier events [72] like pathogen extinction. The distribution of the uncertainty is an assumption [73]. Note that the forecasting models (to be described) are also statistical. The distinction is that, like the deterministic models, the stochastic models simulate a theory to prophesize the future while the forecasting models extrapolate the past into the future.

Graph based epidemiological models can be interpreted as an abstraction of an explanation (or a translation) to a branch of mathematics – graph theory [74, 75] – before *in silico* simulation [76–79]. The latter interpretation provides the flexibility of graph theory or the heritage of an established application like social network theory [80–83]. Here graph or network based methods are therefore classified as orthodox predictive methods and ODE alternatives.

The unorthodox predictive methods also presume that phenomena can be explained but the explanation is not translated into mathematics before simulation. Rather, direct *in silico* simulation of the explanation is performed. Thus some graph based implementations can be interpreted as unorthodox [17, 84–87]. Graphs consist of vertices and edges where (for infectious diseases and social networks) the vertices represent individuals and the edges represent relationships or interactions. Traditionally, the vertices have no geometric interpretation and do not simulate spatial spread [88] but the vertices can be mapped to location [86].

Agent based models (ABM) [6, 16, 89, 90] and cellular automata (CA) [91–93] are spatial, unorthodox, predictive models and PDE alternatives. CA are constructed on a regular lattice and this restriction is removed for ABM [94]. As examples of Artificial Life [95], an agent (or node) acts independently subject to simple rules on the local environment. The collective can prophesize complex phenomena that other predictive methods cannot [96]. These models simulate heterogeneity and mixing [97] but the PDEs that they represent are not apparent [43, 98]. CA can reduce to ODEs [99] and for at least one application (computational fluid dynamics) the PDEs that they represent have been derived [100]. Lattice gas cellular automata (LGCA) [99] and probabilistic(PCA) or stochastic cellular automata (SCA) [92, 93] are subclassifications of CA [43, 98].

Forecasting presumes that phenomena have a recognisable and reproducible pattern. Forecasting fits a curve to a historical pattern and extrapolates the pattern into the foreseeable future [55, 101]. The Fourier theorem states that any curve can be reproduced by an infinite series of superimposed sinusoidal waves [23,102–106]. Filtering refers to the attenuation (or omission) of frequencies that do not substantively contribute to the signal [23, 102, 107] – resulting in a finite series. In electrical engineering, signal noise is presumed to have high frequency. A low-pass filter (allowing low frequencies to pass) attenuates the noise and smooths the resultant signal [102]. Generally, smoothing is a subset of filtering [102] that attenuates high frequency signals.

The Box-Jenkins forecasting models [108, 109] also fit curves. The prototype is the autoregressive moving average (ARMA) model that forecasts weakly stationary behaviour. The autoregressive integrated moving average (ARIMA) includes trend by differencing to transform the ARIMA model into a stationary model [108–110]. Seasonality (periodicity) can also be incorporated into these time series models [109]. The term autoregression refers to historical data points of a curve being used to estimate the model parameters that predict future values of that same curve [111]. Autocorrelation is a metric of how well past results may foretell future results.

ARIMA models fit a linear combination of a finite number of earlier observations and their differences – parsimonious models [108]. See Mills [111] chapters 6 and 11 for an introduction to non-linear functions. The curve fitting distinguishes the traditional time series models [18, 112, 113] from the artificial intelligence (AI) time series models [112]. Traditional, statistical estimation methods include the maximum likelihood method, the conditional sum of least-squares and the ordinary sum of least-squares [108, 109, 114, 115]. AI is an umbrella term for a collection of methods that searches a space for an adequate solution. In epidemiology, the AI methods search for parameter combinations that result in adequate curve fitting [19, 116, 117]. Although the parameters are not necessarily optimal, AI excels at non-linear models with or without *a priori* knowledge or understanding of the system’s behaviour [118].

State-Space models are a subset of signal-plus-noise problems [107] and are introduced as a form of forecasting [109, 111]. Briefly, an observation (space) equation and a state equation are coupled. Each of these equations has a superimposed uncertainty that is assumed Gaussian [119]. The observation (measurement) equation’s independent variable is the signal. In infectious epidemiology; reported new cases, disease mortality [120], waste water serology [121] and combinations thereof are examples of signals. The signal can be a proxy [122, 123] that can be affected by both testing strategy and implementation [120]. For example, South Korea’s strategy of significantly increasing access to testing [124] in COVID-19, may have affected the signal quality. Conceivably, universal testing is more effective [125, 126] but less efficient [127] than opportunistic, symptomatic testing [128–131]. Nevertheless, these strategies should converge when asymptomatic infection is rare. Conceivably, a well implemented track-and-trace policy can outperform a poorly promoted/implemented universal testing policy [127, 132, 133].

The unobserved state function is based on *a priori* knowledge of a system’s behaviour and can be deterministic [134, 135] or empiric [136, 137]. The measured observation/signal is coupled to an unknown state. Backward and forward recursion approximates a state that corresponds to the signal [107]. The Kalman Filter is a popular recursion method for implementing state-space models [121, 134–139].

The above models require local, disease-specific data for prophesy. Thus, the disease must manifest in that region to determine the model parameters for that region. Cardoso and Gonçalves [21] propose a form for a universal population-size *or* -density dependent scaling law and use regression [101] to determine the parameters for COVID-19. Their approach potentially circumvents the need to determine local modelling parameters locally. Rather, parameters from other centres can be projected – adjusted for local conditions [25]. Figure 2 illustrates the 1 week delay [140] in the stage of spread of the ancestral SARS-CoV2 between the UK and the USA [141]. Given the time dependence of intervention, the universal scaling law may prove more beneficial to regions less connected to the epicentre like India in the COVID-19 example of Figure 2.

**Fig. 2.**
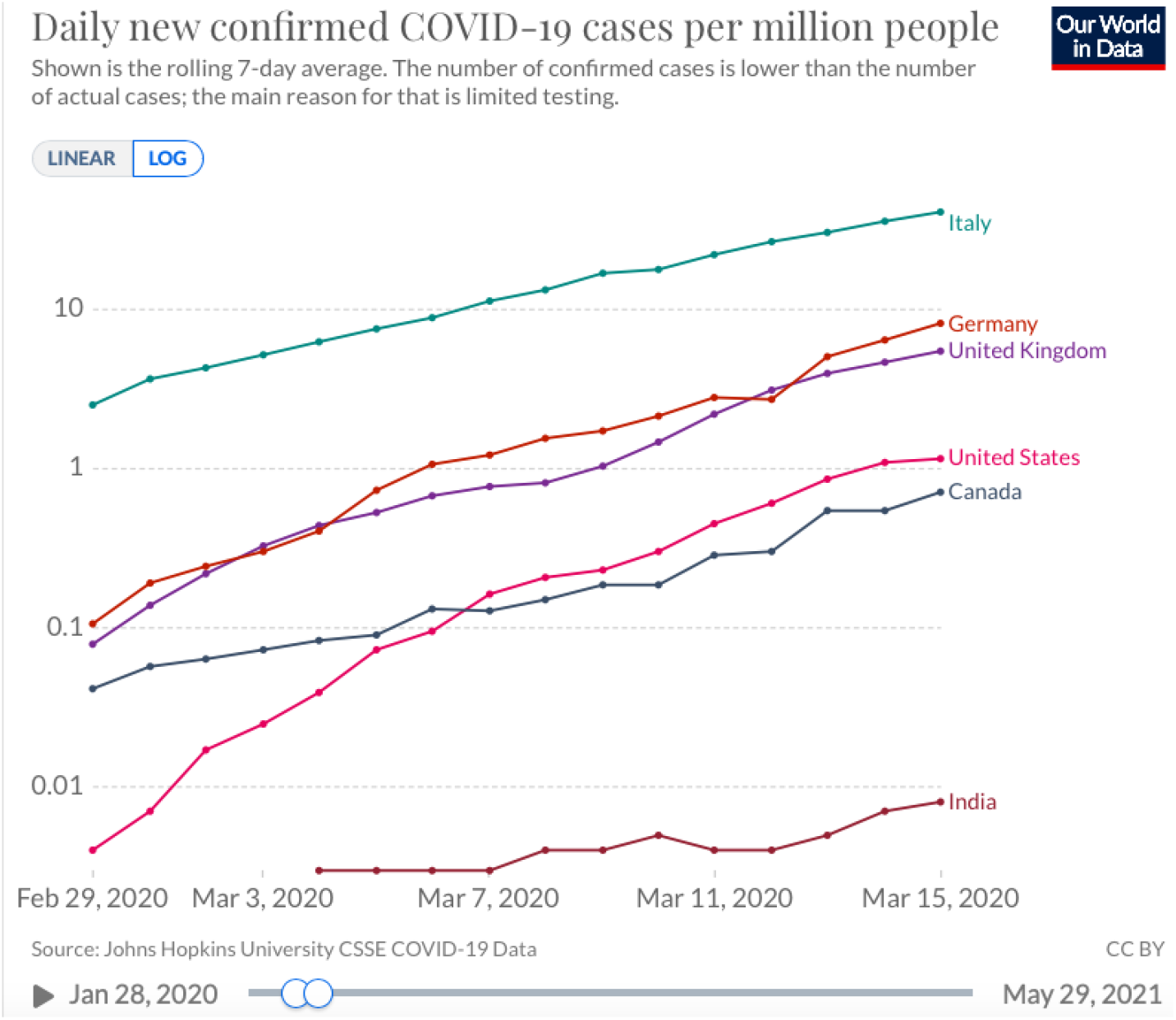
Delay in confirmed cases per million population by country [141]

Hu *et al* [142] potentially circumvent regression’s need for many pre-existing disease centres [21,25] by repurposing formulae from the kinetic theory of ideal gases to derive population-density dependent contact rates. Hu’s contact rates are an alternative to the HI-STR’s law of mass action. The HI-STR prototypes’s contact rate is population-size *and* -density dependent [43].

## 2 Methods

The HI-STR prototype is based on the SIR model but replaces 2 assumptions and is formulated for an isolated population on a surface [43]. Thus

1. it is explicit that the model only applies to sufficiently isolated populations,
2. population density is incorporated because it is formulated on a surface and
3. the PDE model formulation problem is replaced by the problem of defining sufficiently isolated populationss (SIPs) and sufficient behavioural similarity

Hamer’s mass action law [143] assumption is replaced with the law of mass action [43] – its chemistry precursor [144, 145] – such that the probability density function for a single successful transmission is

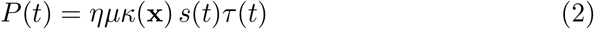

where *η* is an infectious disease-specific variable that reflects avidity, *μ* is a function of mode of transmission, *κ*(**x**) is a function of social behaviour, *s*(*t*) is the density of susceptible individuals and *τ* (*t*) is the density of hosts capable of transmitting the pathogen. The total transmissions (including those of secondary hosts) in a population of size *N ≫*1 and population density *ρ*_*n*_ over the period that the primary host is transmission capable 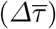 is shown to be

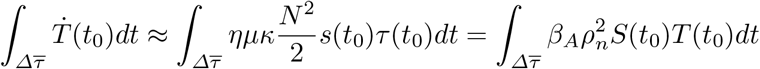

where *S*(*t*) is the size of the susceptible population, *T* (*t*) is the size of the transmission-capable population and 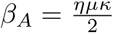 [43].

The SIR prototype’s exponential infectious period assumption is replaced with the HI-STR model’s more biologically appropriate constant transmission period. This results in the SIR-like DDE model.

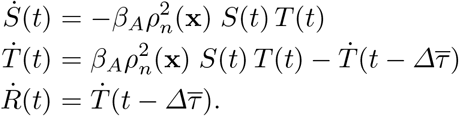

Selecting a timescale (the transmissible timescale) where a time unit (*Δt*) equates to 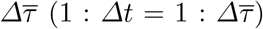 renders the delay negligible – reducing the above DDE to an ODE [43].

Exploiting the periodicity of infection opportunity, a second timescale (the rhythmic timescale) is defined as 1 : *Δt* = 1 : *δt* where *δt* is the period of the infection opportunity cycle. For respiratory infectious diseases, *δt* is the host’s sleep-wake cycle (*ie* 1 day). Given that there are ℬ units of *δt* in 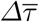, the number of infections after ℬ*δt* is the same as after 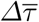. A binomial expansion is used to show that the ODE in the transmissible timescale reduces to

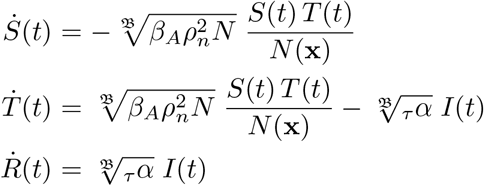

in the rhythmic timescale [43] where _*τ*_ *α* is the infection frequency in the transmissible timescale. The HI-STR model’s rhythmic timescale basic reproduction number for SIP *z* is then [43, 146]

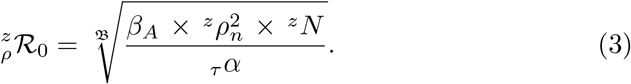

Both *β*_*A*_ and _*τ*_ *α* are dependent on behavioural characteristics that may be cultural [48, 147]. These are assumed constant for behaviourally-similar populations. There is a subtle difference between Böckh’s *ℛ* _*0*_ and its rhythmic timescale equivalent, _*ρ*_*ℛ*_*0*_, but these are used interchangeably here [43].

Dividing Equation 3 for SIP *z* by the same for behaviourally-similar SIP *y* derives Equation 1. It is assumed that the anglophone United Kingdom (UK) and United States of America (USA) have similar concepts of personal space and familiarity with an associated hierarchy of physical interaction rituals [148] such that Equation 1 applies. A metric for behavioural/cultural similarity was not identified. Conceivably, host behaviour could be sufficiently similar across all SIPs. If host behaviour is sufficiently similar across SIPs, then Equation 1 is a universal scaling law (independent of behaviour) and should be compared with Cardoso and Gonçalves’ universal scaling law [21] obtained by regression. The UK and USA were selected to increase the likelihood of a successful validation. From Figure 2, projection would have given the most connected states one week’s lead time. The ancestral SARS-CoV2 pathogen was selected because transmission dynamics data were available, there was no interference from VOCs and the Imperial College London (ICL) group estimated ℛ_0_ for the ancestral SARS-CoV2 in both the USA and the UK..

The same field(reported mortality) and estimation methods [140] were used by the ICL group to measure the ancestral SARS-CoV2’s ℛ_0_ for the UK [140] and the individual states of the USA [149]. Consequently, using these studies to validate the projection of the UK’s ℛ_0_ on to the USA’s states avoids biases introduced by different fieldand estimation methods. Their semi-mechanistic Bayesian hierarchical model is sensitive to the generation interval [140] but a gamma distribution with mode 6.5 days was used for their 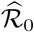 for both the UK and the USA states. From Equation 1, the projection onto state *z* is

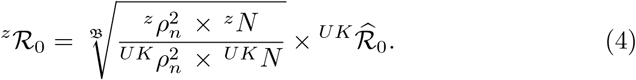

The paired student-t test is used to compare the USA ℛ_0_ estimates 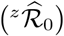 [149] to the UK’s projection on *z (z* ℛ _0_).

Statistical analysis was conducted in the open-source R Project for statistical computing (https://www.r-project.org)

## 3 Results

The transmission dynamics, distribution of pathology, case fatality rate and other clinical, pathological and epidemiological characteristics associated the ancestral (wild type) SARS-CoV2 are collectively designated COVID-19(wt). Appendix 6.1 demonstrates that the transmissible timescale for COVID-19(wt) is 1 : 9 days and the rhythmic timescale is 1 : 1 day. The ratio of the time units in these timescales (ℬ) is 9.

The UK’s estimated ℛ_0_ is 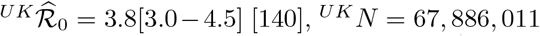, ^*UK*^*N* = 67, 886, 011 with ^*UK*^*ρ*_*n*_ = 280.6 *km*^*−*2^ in 2020 [150]. Equation 4 projects this 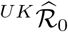 onto the states of the USA adjusting for each state’s population-size and -density. Appendix 6.2 removes any outliers. Figure 3 compares the 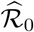 density distribution [149] of the remaining 40 states to those projected from the UK estimate 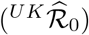 [140]. Figure 3(a) projects the median UK estimate 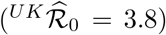 while Figure 3(b) projects 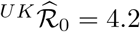. The latter remains within the uncertainty of the UK’s *R* 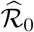 estimate [140].

**Fig. 3.**
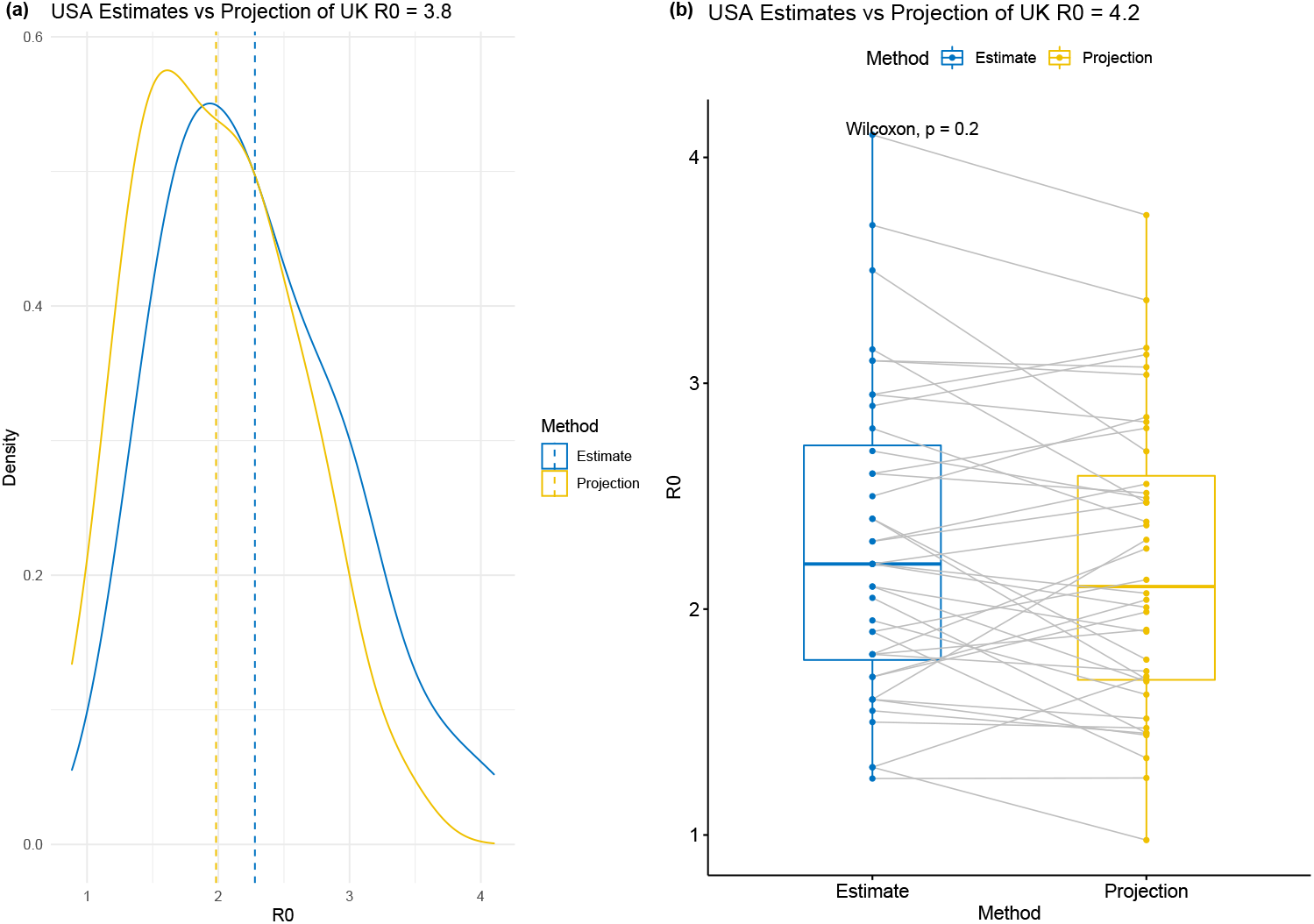
(a) Density distribution comparison between estimated ℛ_0_ the median estimated ℛ_0_ for the UK projected on to the USA’s state (b) Box-and-whisker plot comparison between estimated ℛ_0_ and a UK 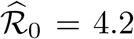 projected on to the USA’s states

Table 1 summarises the results of the paired student t-test comparing the estimated ℛ_0_ [149] of the 40 USA states 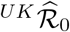 to the projections of the 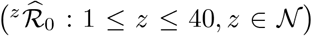 (within the range [3.0 4.5]) [140] on to those 40 states.

**Table 1.** Comparison of paired student *t* -test results between estimated and projected basic reproduction numbers in the USA for various UK basic reproduction number estimates. *μ*_*d*_ is mean of differences. *CI*_*d*_ is 95% confidence interval of the differences. *N* = 40.

| | Estimated UK basic reproduction number ($^{UK}\hat{\mathcal{R}}_0$) | | | | | |
| --- | --- | --- | --- | --- | --- | --- |
| $^{UK}\hat{\mathcal{R}}_0$ | 3.0 | 3.4 | 3.8 | 4.1 | 4.2 | 4.5 |
| $p$ | $7 \times 10^{-15}$ | $5 \times 10^{-11}$ | $4 \times 10^{-6}$ | 0.02 | 0.13 | 0.24 |
| $\mu_d$ | 0.7 | 0.5 | 0.3 | 0.1 | 0.1 | -0.1 |
| $CI_d$ | [0.6, 0.8] | [0.4, 0.6] | [0.2, 0.4] | [0.0, 0.25] | [0.0, 0.2] | [-0.2, 0] |

Of note, for 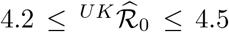, a statistically significant difference between the estimated and projected ℛ_0_s does not exist for those 40 states. For 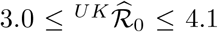, although there is a statistically significant difference between the estimated and projected ℛ_0_; this difference is not epidemiologically significant when compared to the uncertainty in 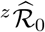 for those states [149].

The model assumes no inherent immunity. Furthermore, the parameter estimates for ℬ have considerable variation (Tables 2 and 3). Appendix 6.3 is a sensitivity analysis demonstrating that up to an inherently immune fraction of 50% the change in the ^*z*^ℛ_0_ projections is not significant compared to the uncertainty in 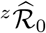 – Figure 4(b). Similarly, changing the symptomatic fraction does not cause an epidemiologically significant change in ℛ_0_ – Figure 4(a). The infectious and transmissible periods are varied in Figures 4(c) and (d), respectively.

**Table 2.** Asymptomatic prevalence for COVID-19(wt).

| $n$ | Infected if tested | Asymptomatic | Population |
| --- | --- | --- | --- |
| 36 | 53% | 11% | Human challenge trial [199] |
| 3711 |  | 17.9% | Diamond Princess Cruise Ship [200] |
| 565 | 2.3 % | 30.8 % | Japanese evacuees from Wuhan [193] |
| 10090 |  | 31 % | Seven whole population meta-analyses including Vo’, Italy [198] |
| 5155 | 2.0 % | 42.2 % | Vo’, Italy [201] |
| 3711 | 19.2 % | 46.5 % | Diamond Princess Cruise Ship [202] |
| 1766 | 59.4 % | 47.8 % | Charles de Gaulle Aircraft Carrier [195] |
| 4954 | 17.3 % | 58.4 % | USS Theodore Roosevelt [195] |
| 217 | 59 % | 81.3 % | Cruise Ship Ernest Shackleton [131] |

**Table 3.**
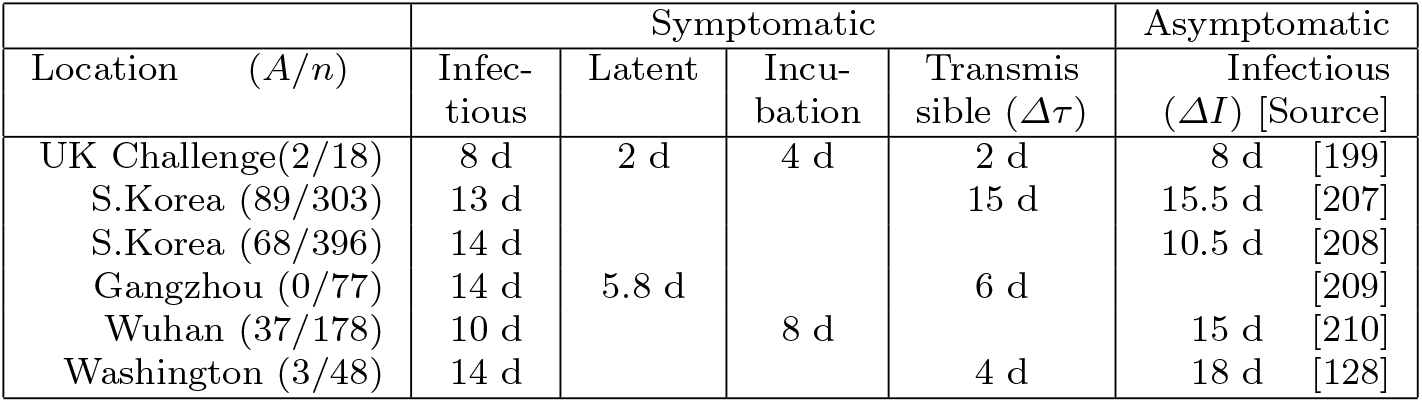
Median latent, incubation, infectious and transmissible periods for symptomatic and asymptomatic COVID-19(wt) patients from early 2020. *A* = *asymptomatic, d* = *days*.

**Fig. 4.**
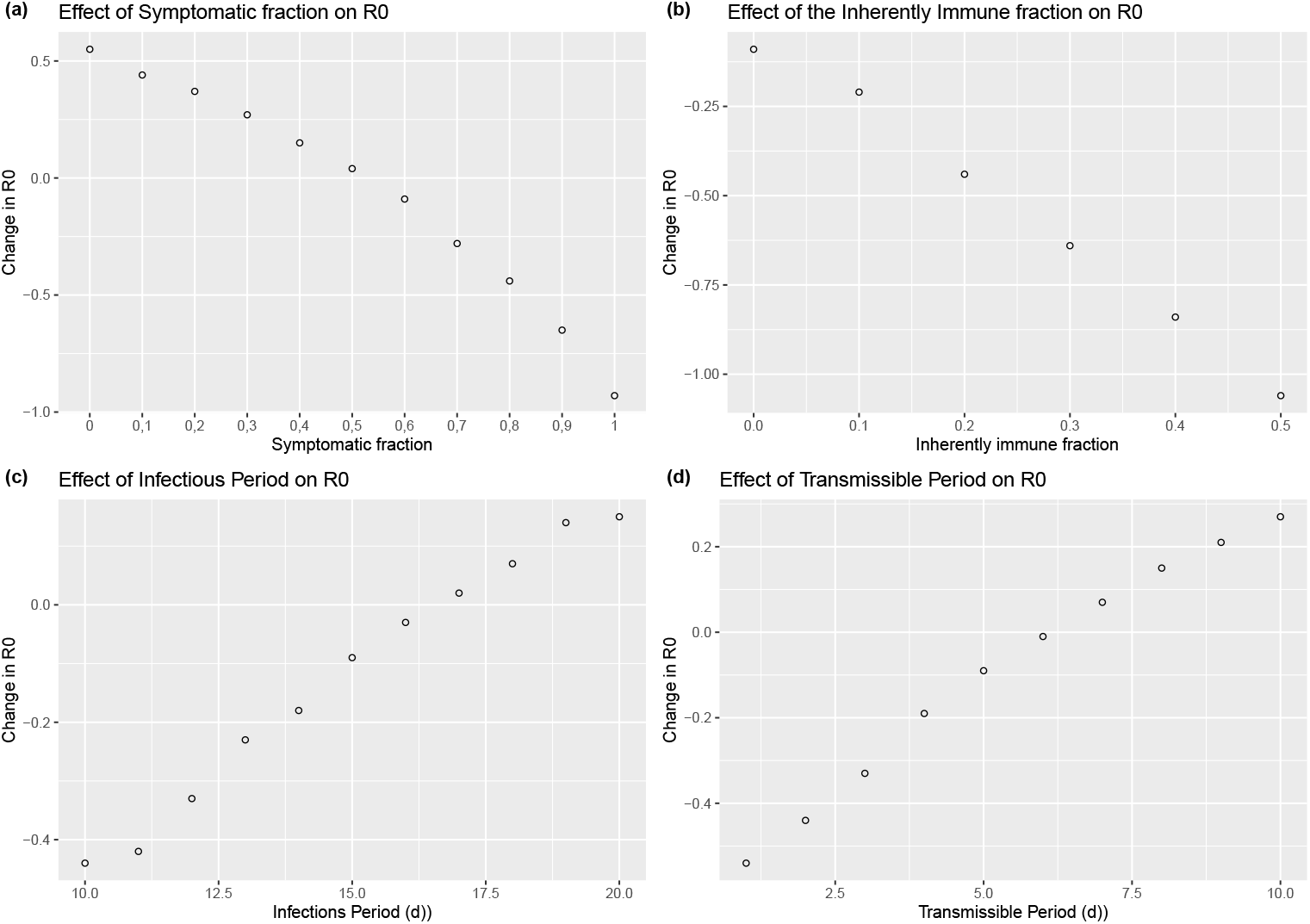
Sensitivity Analysis for COVID19(wt): (a) Symptomatic fraction (b) Inherently immune fraction (c) Infectious period (d) Transmissible period

Despite the above, the HI-STR predicts that increasing the symptomatic fraction decreases ℬ and, consequently, ℛ_0_ by increasing the contribution of those with a shorter transmissible period (Figure 4(a)). It confirms that increasing the inherently immune fraction reduces reduces ℛ_0_ (Figure 4(b)). As expected, increasing the infectious or transmissible periods increase ℬ and therefore ℛ_0_ (Figures 4(c) and (d)).

## 4 Discussion

Coronavirus disease 2019 (COVID-19) is the umbrella term for the diverse pathological manifestations of severe acute respiratory syndrome coronavirus 2 (SARS-CoV2) and its variants. New variants have the potential to supplant pre-existing variants. Projection provides an efficient method to prophesize variant-specific resource requirements. The hybrid incidence, susceptibletransmissible-removed (HI-STR) has demonstrated that projection can be used to foretell the impact of a pathogen variant (the ancestral SARS-CoV2) on the individual states of the United States of America (USA) provided an estimate exists for the United Kingdom (UK). This was possible because the HI-STR accounts for the effect of population characteristics on the basic reproduction number (ℛ_0_). These regions were selected because it is assumed that the individual states of the USA are sufficiently isolated from each other and because it is assumed that these anglophone regions have sufficient behavioural similarity.

It should be noted that the HI-STR prototype does not include the effect of demography [151] on _0_ estimation and projection but age-stratified SIR models can be adapted for the HI-STR. Genetically or behaviourally predisposed individuals also represent subpopulations that affect the average transmission period. For COVID-19, diabetics are a subpopulation that are at increased risk of severe disease and death [152, 153]. The prototype does not include the effect of the distribution of predisposed subpopulations. Clearly the individual states of the USA are not homogenous but a common set of acceptable public behaviourial norms must exist.

Hawaii, Montana, Alaska and Wyoming are among the outliers. The HISTR projections over-estimate ℛ_0_ for these states. For Hawaii, the sea acts as a natural barrier between SIPs. Because the HI-STR is non-linear, these regions cannot be combined. Combining SIPs artificially increases ℛ_0_ projections. Communities within Alaska, Montana and Wyoming may be sufficiently isolated for them to be treated as SIPs. Conversely, states like New York and Washington, DC may not be sufficiently isolated. Neither the method for aver-aging ℛ_0_ estimates across SIPs nor the estimation of *R*_0_ across SIPs is obvious.

This work’s motivation is the timeous preparation for the local impact of novel pathogen or new VOC. Implicitly, each variant is being treated as a new pathogen to which the local population is completely susceptible. The SARS-CoV2 variants are sufficiently closely related that both vaccination and previous infection by the incumbent may confer immunity to the new variant in some individuals. Thus the projection represents an upperbound in which the challenger VOC replaces the incumbent [42]. This model does not address an equilibrium states where VOCs form a mixture [34]. Intuitively and theoretically, the inherently and naturally immune individuals should affect the transmission dynamics of the variant and the transmissible period. Given the uncertainty in the wild type COVID-19 ℛ_0_ estimates, here it was not possible to show that these would have an epidemiologically significant impact.

Intuitively, asymptomatic carriers increase the reproduction number [154]. Uniquely, the HI-STR predicts this phenomenon (See Appendix 6.3) but, given the uncertainty in the SARS-CoV2 ancestral ℛ_0_ estimates, this effect is not epidemiologically significant. For diseases where a correlation exists between symptoms and mortality, an intervention that only converts symptomatic individuals into asymptomatic individuals may reduce mortality. Ironically, the theory predicts that such an intervention will increase ℛ_0_.

## 5 Conclusion

When confronting a novel pathogen, the impact of the disease has to be foretold to prepare accordingly. Some of these impacts are the basic reproduction number (a proxy for how fast the disease will spread), mortality and morbidity. Some of the impacts that are beyond the scope of this document are the economic and socio-political instability caused by the disease and the intervention.

The hybrid incidence, susceptible-transmissible-removed (HI-STR) prototype is a deterministic alternative to the susceptible-infectious-removed prototype and its ordinary differential equation (ODE) model derivatives. In principle, it has two advantages over these more mature models – it incorporates population-size and -density in the model, and it includes a social or behavioural component. The latter is controversial but it should be noted that the HI-STR has the flexibility to include a social component in the model. It may be that physical interaction across regions and cultures is sufficiently similar (from an infectious disease perspective) that this variable can be treated as a constant. In the latter case, the HI-STR derives a populations-size *and* -density dependent universal scaling law for ℛ_0_.

The importance of the capacity to project a ℛ_0_ for a region is that it allows planning and pre-emptive resource allocation. It also provides a locationspecific ℛ_0_ (baseline) to evaluate interventions. This manuscript demonstrates that the HI-STR model can project the UK’s ancestral SARS-CoV2’s ℛ_0_ onto the states of the United States. It must still be demonstrated for other anglophone and non-anglophone regions.

There are 2 parts to the intervention – the intervention policy and the policy implementation. Policies and strategies [155] can only be evaluated retrospectively [156–160] because of unforeseen long-term risks [161–166] and unintended consequences [167–171]. For regions with the same intervention policy, the location-specific ℛ_0_ provides a baseline to compare intervention implementation across those regions.

A weakness that the HI-STR currently shares with the other ODE models is that it does not predict the waves of infection seen with both SARS-CoV2 and the Spanish influenza of 1918 [40]. Models can be constructed with periodic interventions or behaviour resulting in periodic infections [29]. SARS-CoV2 has demonstrated that some of these waves may be due to new variants outcompeting incumbents [29]. The HI-STR model does not account for pathogen evolution and random events like VOCs but here it has been demonstrated that such an event can still be projected timeously. Seasonal changes in pathogen biology or behaviour can be incorporated into the avidity term (*η*) and seasonal changes in host behaviour can be included in the social behaviour term (*κ*(**x**)) of the transmission probability density function (Equation 2). Migration requires a spatial model [40].

The HI-STR predicts that an intervention that only converts symptomatic individuals into asymptomatic individuals will increase ℛ_0_. A risk that can therefore be foreseen is that, even if the virion mutation rate remains constant [35], an increased ℛ_0_ should increase the pathogen population’s mutation rate.

## 6 Appendices

### 6.1 The HI-STR parameters for the ancestral SARS-CoV2

COVID-19 is a collection of clinical symptoms and pathologies [172, 173] assigned to several variants of SARS-CoV2 [174–176]. The heterogeneity of pathology [177, 178] is due to both variable host responses to a variant [179– 182], and multiple viral lineages [183, 184], their variants [185, 186] and their mutations [187, 188]. Here COVID-19(wt) will refer to the distribution of pathology, case fatality rate, severity and transmission dynamics of the subset of COVID-19 due to L lineage of the ancestral (wild type) SARS-CoV2 [181] to distinguish it from the corresponding findings of the *α* (B.1.1.7) [189], *β* (B.1.351), *δ* (B.1.617.2) [36, 183] and *o* (B.1.1.529) [186] variants.

Benjamin [43] defines a time unit in the transmissible timescale as the weighted average of the time a host can transmit a pathogen. The transmission period is limited by viral load in a latent period, recovery, death, pharmacological intervention and behavioural adaptation like quarantine. The weighting is based on the relative proportions of inherently immune (1 *− σ*), symptomatic (*ψ*), asymptomatic (1 *− ψ*), and other subpopulations with distinct transmis-sion periods.

The assumption is that, for a novel pathogen, when sufficient contact is made between a host and a potential host (in a completely susceptible population) there are 3 possible outcomes. There are inherently immune/resistant individuals (not previously exposed) that will not become infected and therefore have a transmission period of zero [190–192], there are asymptomatic individuals [129,193] that may have the transmission period shortened by clearing the virus [189,194] or prolonged by not isolating [53,195], and the symptomatically infected who will have the transmission shortened by either self-isolation, hospitalisation or death. In principle, each of these group’s transmission periods and transmissibility [194, 196] are dependent on the demography [151, 195],comorbidities [152, 153] and genetic predispositions [197] within that group.

The prevalence of asymptomatic infection has been reviewed [195, 198]. Whole population survey’s from 2020 of presumed COVID-19(wt) and a young adult challenge trial [199] are presented in Table 2. The prevalences may re-flect demography. The median asymptomatic prevalence of 42.2% will be used further.

The infectious period and infectiousness of these groups have been reviewed [61, 196, 203, 204]. Each study uses viral load as an imperfect proxy for infectiousness [128, 204–206]. Lavezzo *et al* [201] shortens the viral particle shedding period by 4 days [128,206] to determine the infectious period because the host continues to shed inactive viral RNA for 4 days before the RT-PCR assays can no longer detect them. Table 3 presents the latent, incubation and infectious periods for the symptomatic and asymptomatic.

The latent period is from infection to sufficient viral shedding for successful transmission. Incubation is from infection to symptom onset. The period of time that a virus is detectable has been shortened by 4 days to obtain the infectious period because of the detectable inactive viral RNA post recovery.. It is assumed that symptomatic patients self-isolate, are hospitalised or ostracised at symptom onset or within a day thereof. Therefore the transmissible period is the difference between the incubation and latent periods.

In each of these studies there is no demonstrable difference in infectivity between these groups [203, 204, 206, 208]. Table 3 mitigates the bias of retrospective studies [194] by only including whole population studies. Nevertheless each population is unique and not necessarily representative. Of note; there may be small differences in the definition of symptomatic and asymptomatic individuals, different real time reverse transcriptase polymerase chain reaction (RT-PCR) platforms were used and different cycle count thresholds used.

The proportion of naturally immune or resistant (1*− σ*) is unknown. Although the UK challenge trial [199] is prospective, neither the sample nor the inoculum are necessarily representative. It is assumed that the naturally immune and resistant are negligible (*σ* = 1). In summary, 42.2% are asymptomatic carriers with a 15 day transmission period (*Δτ*) and the symptomatic 57.8% have *Δτ* = 5 days. Consequently, the weighted average transmission period is *Δτ* = 9 days and the transmissible times scale is 1 : 9 days.

Periodic human behaviour is the result of superimposed daily, weekly, monthly and annual cycles. For infectious diseases, the rhythmic timescale is determined by the periodicity of transmission opportunity. For airborne diseases like COVID-19, the dominant cycle is diurnal with maximum transmission opportunity during day time social interactions and reduces to a minimum while sleeping. The periodic timescale (1 : *δt*) for COVID-19 is thus 1 : 1 day.

The ℬ constant in Equations 1 and 3 is the ratio of a unit of time in transmissible times scale 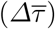 to a time unit in the rhythmic timescale (*δt*). Thus, for COVID-19(wt), ℬ= 9.

### 6.2 States of the USA basic reproduction number outliers

Re-arranging Equation 3, substituting the median reproduction numbers estimates 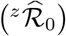 for the 1*≤ z≤* 51 states of the USA [149] and, the population-sizes (^*z*^*N*) and -densities (^*z*^*ρ*_*n*_) for these states [150] allows one to determine the proportionality constant 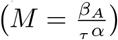 in

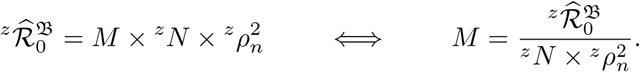

Figure 5 (the density distribution for *M*) identifies 6 outliers. The median

**Fig. 5.**
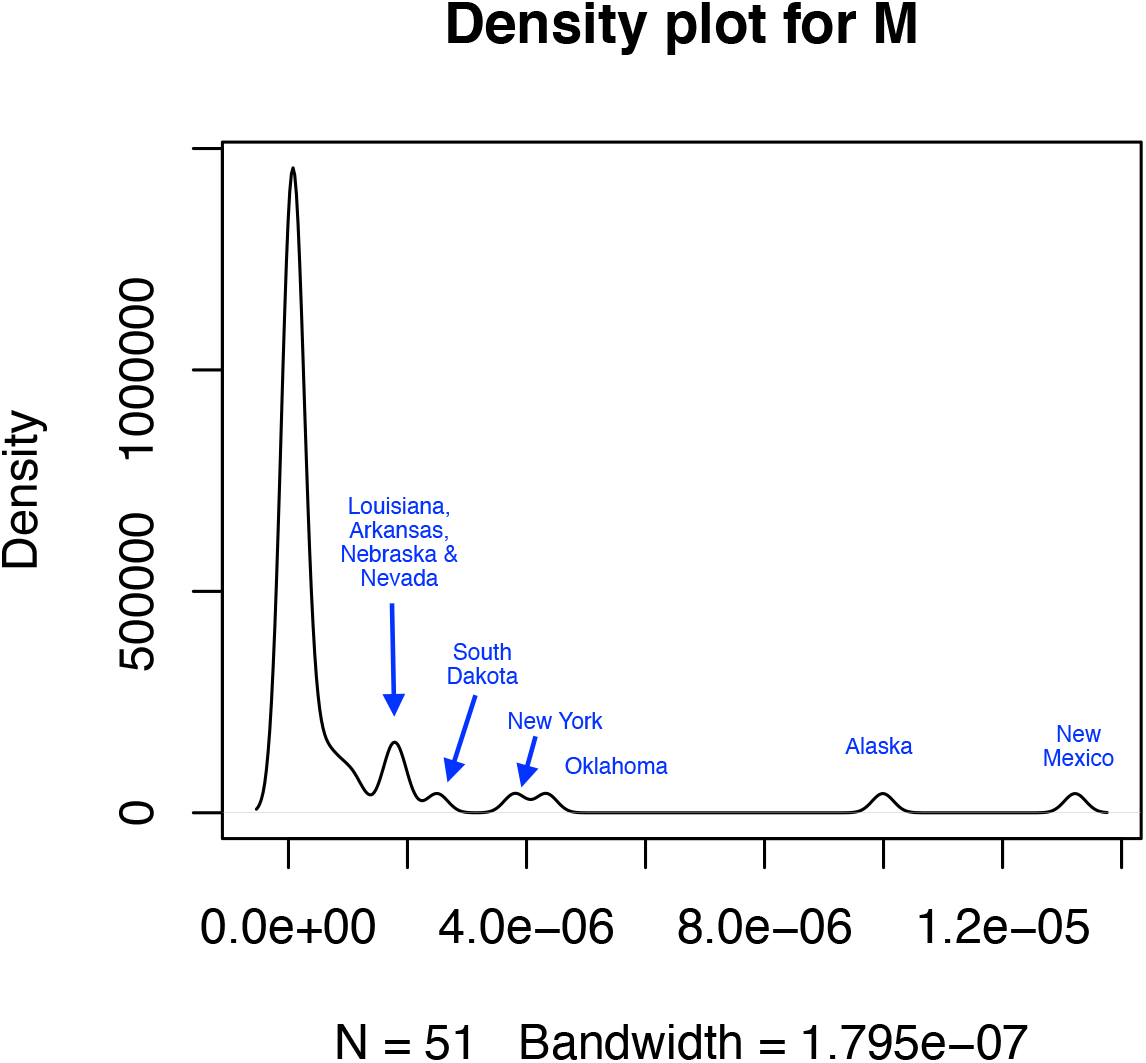
Population density distribution of *M* in 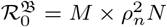

[IQR] of the proportionality constant *M* is 1.1 *×* 10^*−*7^[3.4 *×* 10^*−*8^, 6.2 *×* 10^*−*7^].

New Mexico, Alaska, Oklahoma, New York, South Dakota and Louisiana are removed from further analysis.

The relative error between the predicted basic reproductive (*z* ℛ_0_) and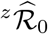 :

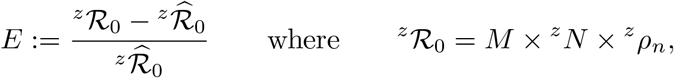

is assumed to have a Gaussian distribution about a mean of 0. The Median [IQR] of *E* is *−*0.02[*−*0.17, 0.05]. Figure 6 is the density distribution of the rela-tive error between the predicted and measured reproductive numbers. Hawaii, Montana, Minnesota, Wyoming and Washington DC are removed as outliers.

**Fig. 6.**
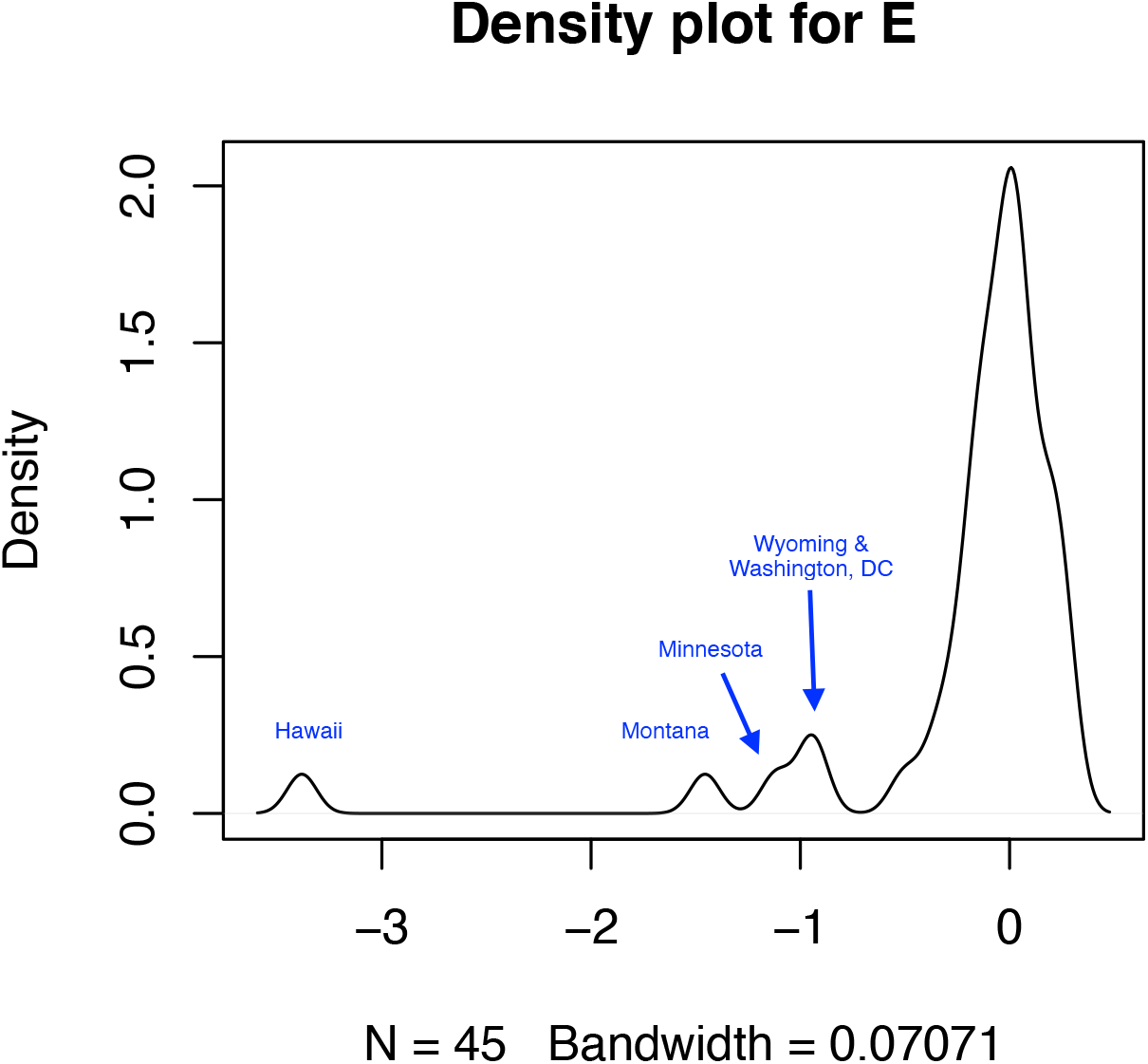
Population density distribution of *E*, the relative error between the observed and predicted basic reproductive numbers

Figure 7 compares the density distributions of 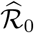 and ℛ_0_ for the remaining 40 states of the USA and demonstrates that the relative error, *E*, is normally distributed. The Shapiro-Wilk test on *E* could not exclude normality (*p* = 0.31).

**Fig. 7.**
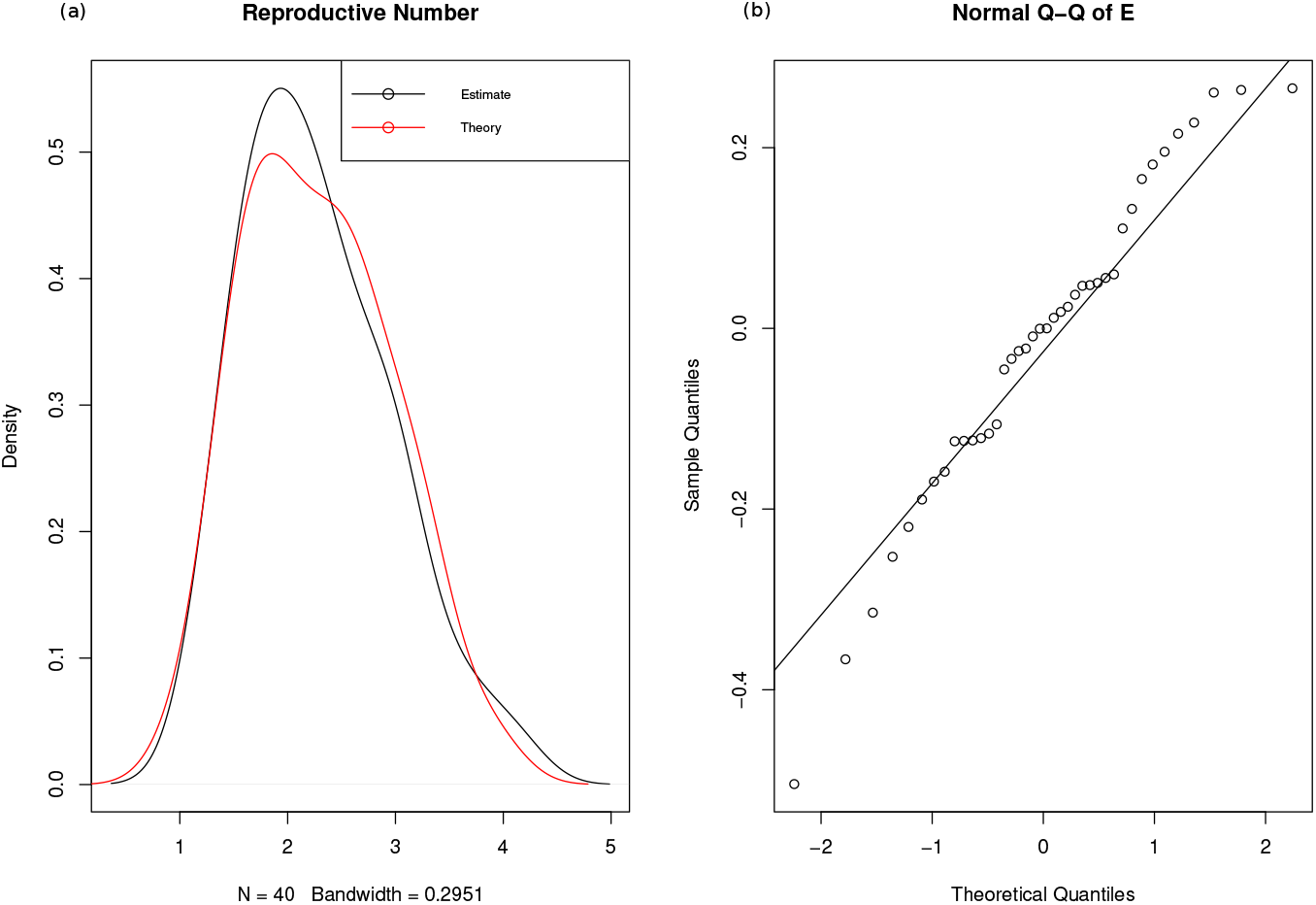
(a) Comparison of the theoretical and estimated _*ρ*_ ℛ_0_, (b) Demonstration of the normality of the relative error *E*

### 6.3 Sensitivity analysis – impact of asymptomatic ratio

The ratio of the time units in the transmissible timescale to the time units in the rhythmic timescale (ℬ) translates to the average transmissible period 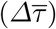 for the special case where the time units in the rhythmic timescale (*δt*) = 1. For this special case, ℬis dependent on 4 variables

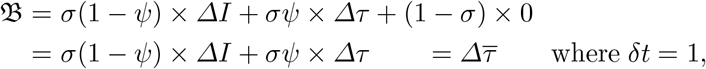

*σ* is the non-immune or susceptible portion of the population, 1 *− σ* is the immune portion that cannot be infected, *ψ* is the portion of *σ* that will be symptomatic if infected, 1 *− ψ* is the proportion that will be asymptomatic if infected, *ΔI* is the infectious period and *Δτ* is the transmissible period.

Thus far, this manuscript has assumed that *σ* = 1 while Tables 2 and 3 demonstrate the considerable variance in the estimates of *ψ* and *ΔI* and *Δτ*, respectively. Figure 4 is a sensitivity analysis depicting the effect of changes in:

a. the symptomatic fraction (*ψ*) when the inherently immune fraction (1 − *σ*), infectious period (*ΔI*) and transmissible period (*Δτ*) are held constant at 0; 15 days and 5 days, respectively;
b. the inherently immune fraction (1*− σ*) is varied from 0 to 0.5 with *ψ* = 0.6, *ΔI* = 15 days and *Δτ* = 5 days. The distribution of the constants *M* and *E* deviate from normality for *ψ >* 0.5;
c. infectious period (*ΔI*) with 1 *− σ* = 0, *ψ* = 0.6 and *Δτ* = 5 days and
d. transmissible period (*Δτ*) with 1 *− σ* = 0, *ψ* = 0.6 and *ΔI* = 15 days

on the difference between UK’s projection of 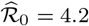 onto the USA states and the USA estimates of ℛ_0_ – projection - estimate.

As expected, increasing the symptomatic fraction (*ψ*) or the inherently immune fraction (1 *− σ*) reduced ℛ_0_. Of note, relative to the estimates and the uncertainties in these estimates [149] the reduction in ℛ_0_ is epidemiologically insignificant over the domains investigated. The *E* distribution deviates from normality on the Shapiro-Wilk’s test for inherently immune ratios greater than 40% and visibly for ratios greater than 50%.

Increasing either the infectious period (*ΔI*) or the transmissible period (*Δτ*) increased ℛ_0_. Again, relative to the estimates and the uncertainty in these estimates [149], the increase in ℛ_0_ is epidemiologically insignificant for wild type COVID19.

## Data Availability

All data produced in the present study are available upon reasonable request to the authors

## 7 Declaration

### Availability of data and material

Not applicable

### Code availability

Not applicable

### Ethics approval

Not applicable

